# Stage-Structured, Distributional Prediction of IVF Outcomes with Conditional Updating

**DOI:** 10.1101/2025.09.27.25336680

**Authors:** Alexander Craig, Laura Wartschinski, Mathew Eyre, Ivan Davidson, Michael Christensen, Tobias Wolfram

## Abstract

**Background:** Current IVF calculators provide either cumulative success probabilities, such as the CDC IVF Success Estimator [1] and OPIS calculators [2, 3, 4, 5], or stage-specific point estimates such as the Orchid Embryo Banking Calculator [6], but they do not quantify uncertainty and cannot incorporate patient-specific outcomes observed during treatment.

**Objective:** To develop a distribution-based framework that (i) produces full probability distributions at each IVF stage and (ii) allows downstream predictions to update when new stage outcomes are known.

**Methods:** We constructed a sequential probabilistic model using fresh, autologous IVF cycles from the Human Fertilisation and Embryology Authority (HFEA) registry (2017–2018) [7] for egg retrieval, maturity, and fertilization, and integrated published clinical studies totaling over 435,000 additional observations for blastocyst formation, euploidy, freeze/thaw survival, and live birth after euploid transfer. Models were validated using 70/30 train-test splits with out-of-sample performance metrics. Egg retrieval is modeled with zero-inflated negative binomial (ZINB) regression; downstream stages apply sequential binomial filters. A “known value selection” mechanism conditionally updates predictions when observed counts are entered.

**Results:** The model generates full probability distributions at each stage of IVF rather than point estimates. Multi-cycle modeling enables comprehensive family planning assessments, while known value updating on combined distributions maintains cycle-specific biology rather than averaging outcomes. When observed values are entered, downstream distributions update accordingly, helping to guide clinical decisions. Out-of-sample validation demonstrates minimal overfitting with train-test R^2^ gaps under 0.007. Distribution evaluation confirms well-calibrated prediction intervals (50% coverage: 50.2%, 80% coverage: 79.2%, 95% coverage: 94.9%). The model is available as a web application at https://www.herasight.com/ivf-calculator.

**Conclusions:** A distribution-based, sequential framework with conditional updating addresses key limitations of existing calculators by providing uncertainty-quantified, stage-aware predictions that adapt to patient-specific outcomes observed during care.

**Study Funding/Competing Interests:** Funded by Herasight Inc. Authors are employees or consultants of Herasight.

## 1 Introduction

In vitro fertilization (IVF) represents a $25 billion global industry serving the 1-in-6 couples affected by infertility [8]. Despite this scale, patients navigate treatment using prediction tools that provide only point estimates without uncertainty bounds, cannot incorporate mid-cycle data, and fail to address the sequential, multi-cycle reality of modern fertility treatment. This gap contributes to treatment discontinuation, with patients often stopping due to psychological burden and unclear expectations rather than poor prognosis [9]. As 92% of patients prefer shared decision-making with physicians, many would benefit from tools that present probabilistic information in more accessible formats [10, 11]. Accurate, personalized predictions empower patients to align expectations with clinical realities, make informed decisions, and maintain hope throughout their path to parenthood [12, 13, 14].

Elective IVF for preimplantation genetic testing for polygenic risk (PGT-P) represents a growing market with distinct prediction needs. PGT-P screens embryos for polygenic conditions including diabetes, heart disease, schizophrenia, and various cancers, with 72% public approval and 81.9% of IVF patients expressing interest [15]. The growth in elective fertility preservation further amplifies demand: 70% of women who froze eggs before age 38 and later thawed at least 20 eggs achieved live births, demonstrating the viability of elective approaches [16]. Critically, elective patients pursuing PGT-P have fundamentally different needs than traditional infertility patients. While infertility patients focus on achieving any pregnancy, elective patients may seek to maximize euploid embryo counts to enable meaningful genetic screening. Increasingly, patients are focused on accumulating multiple euploid embryos for screening and future family building, rather than achieving an immediate pregnancy, a metric that existing calculators’ exclusive focus on pregnancy outcomes fails to address [17, 18]. Further, existing IVF calculators are calibrated on infertility populations with inherently lower success rates, creating systematic underestimation for healthy elective patients. This mismatch underscores the need for prediction models that account for higher baseline fertility and focus on distributional outcomes rather than binary success metrics.

Existing IVF calculators fail to meet these diverse clinical needs. The CDC IVF Success Estimator [1] and OPIS calculators [2, 3, 4, 5] provide only cumulative pregnancy probabilities, failing to capture elective patients’ interest in embryo yield distributions while also lacking stage-specific outcomes or uncertainty quantification. Orchid’s Embryo Banking Calculator [6] decomposes biological stages but lacks confidence intervals, cannot update with observed outcomes, and excludes elective patient profiles. Recent machine learning approaches show promise but remain fragmented to individual steps in the IVF process [19, 20]. The absence of uncertainty quantification undermines clinical trust, as medical AI without confidence bounds compromises patient safety [21, 22]. In IVF, where intermediate outcomes drastically affect final probabilities, point estimates mask critical information.

We present a distribution-based framework treating IVF as sequential stochastic filters with dynamic updating. The model: (1) propagates full probability distributions through biological stages rather than collapsing to point estimates, (2) enables multi-cycle planning while maintaining cycle-specific parameters, and (3) implements conditional updating when observed outcomes become known. Built on 103,924 HFEA cycles [7] with zero-inflated negative binomial egg retrieval, the framework applies sequential binomial transitions calibrated on 435,291 observations [23, 24, 25, 26, 27]. Known value separation maintains cycle-specific biology when updating pooled outcomes.

Our objectives were to develop a sequential distributional model quantifying uncertainty at each IVF stage, implement conditional updating incorporating real-time outcomes, and enable multi-cycle planning preserving cycle-specific characteristics. The web application integration of our model is available at https://www.herasight.com/ivf-calculator.

## 2 Methods

We designed a stage-structured prediction model that treats IVF as a sequence of stochastic filters, propagating full probability distributions through biological transitions: egg retrieval, maturation, fertilization, blastocyst formation, genetic testing, and implantation. Eggs retrieved follow a zero-inflated negative binomial distribution to accommodate excess zeros and overdispersion, consistent with structural zeros (e.g., cancellation/empty follicle) and heavy-tailed egg counts; zero-inflated and negative-binomial models are standard for overdispersed counts. Subsequent stages use binomial filters to propagate distributions, with each stage’s output becoming the input for the next stage (Appendix A.1.3). Success rates for these filters come from our regression models (maturity, fertilization) or age-specific models fitted to published cohorts (blastocyst, euploidy, live birth). Two design choices distinguish our approach: First, we maintain full probability distributions at each stage, quantifying both expected outcomes and uncertainty rather than single point estimates. Second, we implement conditional updating. When an observed value becomes known at any stage, all downstream distributions are recomputed, narrowing confidence intervals and aligning predictions with the evolving clinical course. We developed this framework using a comprehensive dataset combining UK registry data with multiple international clinical studies, as detailed below.

### 2.1 Data Sources

We used the anonymized HFEA 2017–2018 registry [7] for upstream stages (egg retrieval, maturity, fertilization). We restricted to fresh, autologous cycles with valid nonnegative counts and known treatment type, yielding 103,924 cycles for egg retrieval after exclusions. HFEA reports several outcomes in grouped categories (e.g., “6–10 fresh eggs collected”); we applied midpoint imputation, a standard approach for grouped/coarsened data while acknowledging potential bias for open-ended categories (e.g., “*>*40” imputed as 45) [28].

For stages not tracked by HFEA or requiring specialized populations (blastocyst, euploidy, vitrification survival, and live birth per euploid transfer), we synthesized evidence from published studies and national registries totaling over 435,000 additional observations: blastocyst formation from Romanski et al. [23] and Sainte-Rose et al. [24]; euploidy rates from Franasiak et al. [25] and Armstrong et al. [26]; vitrification survival from Gallardo et al. [29] and Coello et al. [27]; and live birth per euploid transfer from SART national registry data. Detailed sample sizes and modeling approaches for each stage are described in their respective subsections.

### 2.2 Statistical Framework

Figure 1 illustrates the complete pipeline and modeling approaches for each stage.

**Figure 1.**
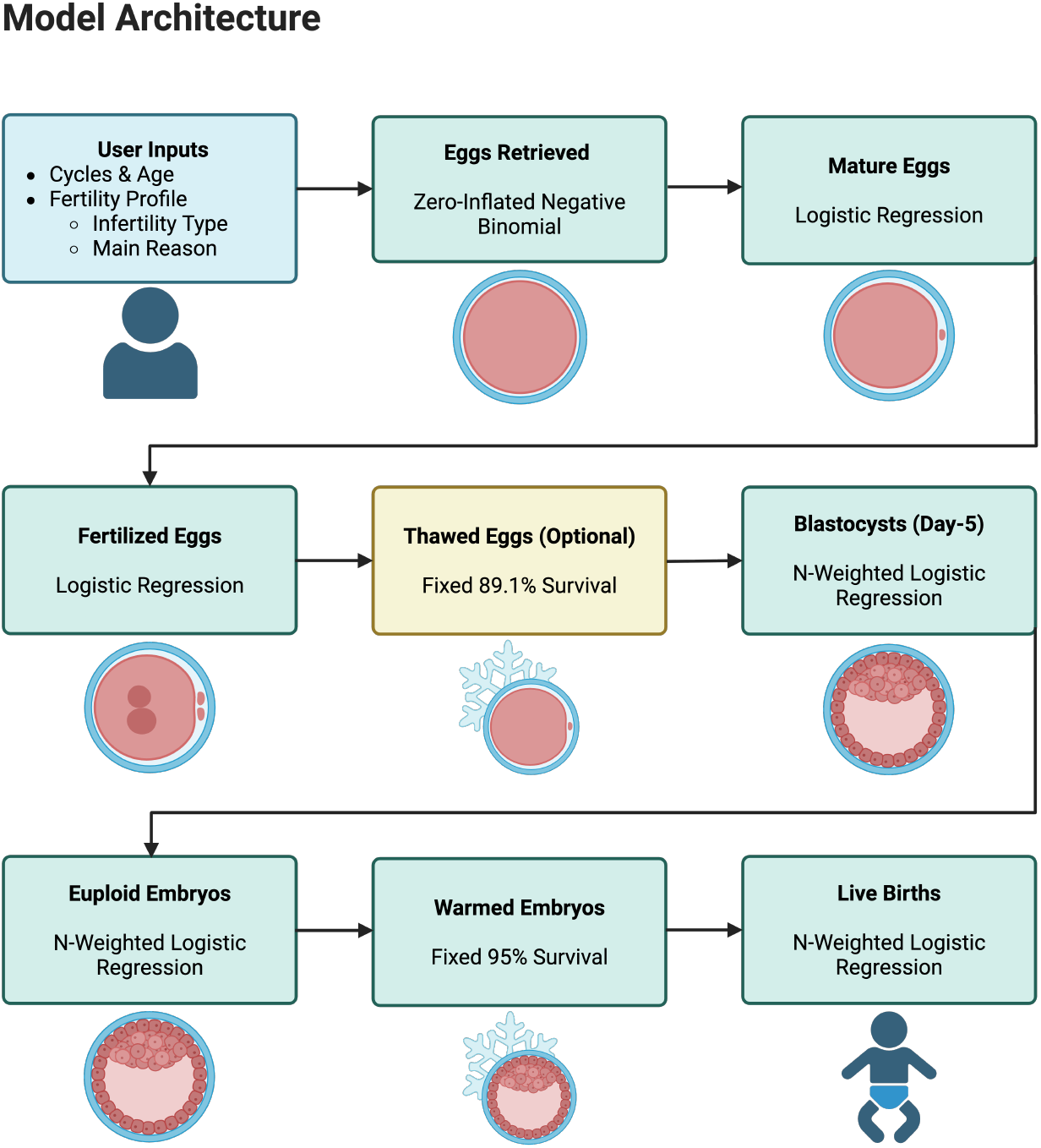
Model architecture showing the stage-structured prediction framework. The model propagates full probability distributions through sequential biological stages, from egg retrieval through live birth. Each stage applies specific statistical models (ZINB for retrieval, logistic regression for transitions, n-weighted logistic for aggregated clinical data) with conditional updating when observed values become known.

The model operates in three interconnected modes:

#### Single-cycle predictions

Each IVF cycle progresses through biological stages with stage-specific success rates. Starting with egg retrieval, the model applies sequential filters representing biological and in vitro processes: maturation, egg thawing, fertilization, blastocyst formation, genetic testing, embryo warming, and implantation. Each stage maintains full probability distributions rather than point estimates, quantifying both expected outcomes and uncertainty.

#### Multi-cycle planning

Patients often undergo multiple IVF cycles at different ages or combine fresh cycles with previously frozen eggs. The model supports heterogeneous multi-cycle scenarios where each cycle maintains independent parameters reflecting its specific age and conditions. For instance, a woman combining 20 frozen eggs from age 32 with a fresh cycle at age 35 benefits from age-specific success rates for each cohort. We combine cycles by discrete convolution to obtain aggregate probabilities while preserving the variance of independent attempts (Appendix A.1.2). This assumes independence across cycles and yields a Poisson-binomial-type total when per-cycle success probabilities differ [30].

#### Dynamic updating with observed values

When actual outcomes become known during treatment, the model updates all downstream predictions conditionally. For single cycles, the observed stage becomes a point mass distribution and subsequent distributions narrow accordingly. For multi-cycle cases, each cycle can specify its own known value at any stage. Cycle distributions are computed independently and then combined through convolution to obtain aggregate probabilities.

### 2.3 Stage Models and Predictors

#### 2.3.1 Fertility Profile System

To simplify user input while capturing clinically meaningful heterogeneity, we developed a fertility profile system that maps seven common clinical scenarios to appropriate model parameters. The HFEA registry predominantly comprises patients with diagnosed infertility, presenting a methodological challenge for modeling the growing population pursuing *elective* IVF, including those seeking preimplantation genetic testing for polygenic risk (PGT-P). To address this, we empirically derived profiles that span the full range of observed outcomes, with special consideration for approximating results in patients without infertility diagnoses.

Each fertility profile configures two categorical variables from the HFEA registry at each IVF stage: the “main reason for treatment” (Treatment-IVF, Egg/Embryo storage, or Donation/Egg share) and the “infertility type” (male factor, endometriosis, ovulatory disorder, tubal factor, or unexplained). The seven profiles are:

- **Non-elective ovulatory**: Treatment-seeking patients with ovulatory disorders, including PCOS
- **Non-elective tubal**: Treatment-seeking patients with tubal factor infertility
- **Non-elective endometriosis**: Treatment-seeking patients with endometriosis
- **Non-elective unexplained**: Treatment-seeking patients with unexplained infertility
- **Non-elective male factor**: Egg/Embryo storage cycles with male factor infertility
- **Elective baseline**: Egg/Embryo storage cycles (conservative scenario)
- **Elective optimistic**: Donation or egg share cycles (optimistic scenario)

The five non-elective profiles represent the traditional IVF population with diagnosed infertility (main reason: Treatment - IVF in the HFEA registry), each maintaining a consistent infertility type across all model stages to reflect a specific diagnosis. Four of these profiles (ovulatory, tubal, endometriosis, unexplained) use “Treatment - IVF” as the main reason in the HFEA registry, directly corresponding to HFEA’s primary patient population. The fifth profile, non-elective male factor, uses “Egg/Embryo storage” as the main reason to better capture egg retrieval patterns, as women with male factor partners often have normal egg retrieval profiles, similar to those pursuing elective storage. Analysis of model coefficients reveals meaningful clinical differences: ovulatory disorders are associated with higher egg retrieval (coefficient +0.316, *p <* 0.001), reflecting the inclusion of polycystic ovary syndrome (PCOS) patients [31], while male factor infertility shows the most pronounced negative impact on fertilization rates (coefficient -0.153, *p <* 0.001).

The elective profiles required special consideration due to the scarcity of true elective cases in the HFEA data. With rising interest in elective IVF for fertility preservation and genetic screening through PGT-P, accurately modeling outcomes for patients without infertility concerns becomes essential. Given the constraints of the HFEA registry, which predominantly captures infertility patients, we developed two profiles using the best available proxies to approximate outcomes for healthy individuals. Notably, the “None” infertility category paradoxically exhibited lower success rates than most diagnosed infertility types, suggesting data quality issues. This category was inferred from an absence of diagnoses rather than explicitly recorded in the HFEA data. We therefore excluded it from our elective profiles despite its apparent relevance.

We developed two profiles by strategically combining infertility category mappings to construct composite elective patient profiles. For each IVF stage, we selected the infertility category that would most closely match patients without fertility issues, based on biological reasoning about which conditions do not affect specific reproductive processes. We chose “Egg/Embryo storage” for the baseline profile as this category captures patients preserving their own eggs or embryos for future use, representing typical elective fertility preservation without selection for exceptional fertility. “Donation/Egg share” suits the optimistic profile because donors undergo rigorous screening for high ovarian reserve and favorable hormone levels, making them specifically selected for high fertility potential [32]. The **elective baseline** profile (main reason: Egg/Embryo storage) uses male factor infertility for egg retrieval, as male infertility does not affect female ovarian response. For maturity and fertilization stages, we use endometriosis, as this condition does not affect oocyte maturation or fertilization processes despite reducing egg quantity [33]. The **elective optimistic** profile (main reason: Donation/Egg share) uses ovulatory disorder for egg retrieval, capturing the higher yields often seen in women with excellent ovarian reserve including those with PCOS [31], then similarly uses endometriosis for maturity and fertilization stages.

This profile system enables clinicians to provide evidence-based guidance for both traditional infertility patients and the growing population pursuing elective IVF, while maintaining model parsimony and clinical interpretability.

#### 2.3.2 Egg retrieval

Egg retrieval presents unique modeling challenges as some cycles result in zero eggs while others show high variability. We used a zero-inflated negative binomial regression model that separately accounts for the probability of retrieval failure and the distribution of positive counts. Predictors include age, fertility profile (capturing main reason for treatment and infertility type), and stimulation status.

##### Training data

The training set comprised 103,924 cycles after applying the exclusions described in Section 2.1.

##### Full versus simplified predictors

A non-parsimonious model evaluated ten predictors: patient age, stimulation status, treatment type (IVF vs ICSI), main reason for treatment, infertility type, previous IVF cycles, previous pregnancies, previous live births, patient ethnicity, and partner ethnicity; the simplified model retained age, main reason for treatment, infertility type, and stimulation status. Reported coefficients below refer to the *count* component.

The parsimonious count model retained: age (coefficient −0.037, *p<*0.001), main reason (treatment vs storage/donation), infertility type (categorical with six levels), and stimulation (coefficient 1.074, *p<*0.001). Main reason coefficients were −0.335 for treatment and −0.103 for storage (vs donation reference). Infertility type showed significant effects with ovulatory (coefficient 0.316, *p<*0.001) having the largest positive impact. Full coefficient tables for all models are provided in Appendix A.2.

Using out-of-sample *R*^2^ on the test set, the simplified model achieved 0.137 (train: 0.140) and the full model 0.153 (train: 0.155); the improvement from simplified to full was 1.6 percentage points.

#### 2.3.3 Regional calibration

Empirical comparison of mean eggs retrieved reveals systematically higher yields in U.S. (SART) data compared to UK (HFEA) data across all age groups, with multiplicative factors ranging from 1.32 to 1.48. This consistent pattern suggests fundamental differences in clinical practice rather. Previous research has documented divergent approaches between U.S. and European IVF practices, including higher gonadotropin doses and different medication protocols in U.S. clinics [34]. These protocol differences likely reflect distinct clinical philosophies: U.S. clinics often prioritize maximizing egg yield given the high financial and emotional cost of repeated cycles, while European practices tend toward milder stimulation approaches. Additionally, differences in regulatory environments, insurance coverage patterns, and patient demographics may contribute to more aggressive stimulation protocols in the United States.

To enable our UK-trained model to provide accurate predictions for U.S. patients, we implemented age-specific multiplicative calibration factors derived empirically from the observed ratios of mean SART to HFEA retrieval rates within each age band. These factors (1.48 for age *<*35, 1.40 for 35-37, 1.36 for 38-40, 1.32 for 41-42, and 1.32 for 43+) scale predictions proportionally while preserving the underlying biological relationships captured by the model. The consistency of elevated U.S. yields across all age groups supports our hypothesis of systematic protocol differences rather than population-level biological variation. While this population-level adjustment represents a pragmatic solution necessitated by the absence of individual protocol data, it enables cross-regional applicability of our framework. Future work incorporating detailed stimulation protocols could refine these calibrations to account for individual treatment intensity.

#### 2.3.4 Maturity and fertilization rates

Maturity and fertilization rates were modeled using logistic regression on HFEA data. Because the registry does not explicitly track metaphase II oocytes, we approximated maturity as the ratio of eggs mixed to eggs retrieved. In standard laboratory workflows, insemination is performed on MII oocytes; thus eggs mixed is a practical proxy for maturity in registry data. Similarly, fertilization was estimated as embryos created divided by eggs mixed, aligning with established laboratory performance indicators. Both models include age, treatment type (IVF vs ICSI), stimulation, and infertility type as predictors.

##### Maturity model training data

The dataset comprised 90,479 cycles after applying relevant exclusions. A 70/30 train-test split was applied with stratification by age.

##### Maturity model full versus simplified predictors

A non-parsimonious model evaluated the same ten predictors as the egg retrieval model; the simplified model retained age, stimulation, treatment type, and infertility type. Reported coefficients below refer to the binomial logit model.

The parsimonious model retained: age (coefficient 0.005, *p<*0.001), stimulation (coefficient ™0.782, *p<*0.001), treatment type (coefficient 2.509 for IVF vs ICSI, *p<*0.001), and infertility type (categorical with male factor coefficient ™0.115, *p<*0.001). See Appendix A.2 for complete coefficient tables.

Using predicted counts against observed mature eggs on the test set, the simplified model achieved *R*^2^ = 0.879 (train: 0.886) and the full model *R*^2^ = 0.879 (train: 0.886); the improvement was 0.02 percentage points. The mean maturity rate was 90.7%.

##### Fertilization model training data

The dataset comprised 90,088 cycles after applying relevant exclusions. A 70/30 train-test split was applied with stratification by age.

##### Fertilization model full versus simplified predictors

A non-parsimonious model evaluated the same ten predictors; the simplified model retained age, treatment type, stimulation, and infertility type. Reported coefficients below refer to the binomial logit model.

The parsimonious model retained: age (coefficient 0.004, *p<*0.001), treatment type (co-efficient ™0.296 for IVF vs ICSI, *p<*0.001), stimulation (coefficient ™0.303, *p<*0.001), and infertility type (categorical with male factor coefficient ™0.153, *p<*0.001). See Appendix A.2 for complete coefficient tables.

Using predicted counts against observed fertilized eggs on the test set, the simplified model achieved *R*^2^ = 0.643 (train: 0.648) and the full model *R*^2^ = 0.643 (train: 0.649); the improvement was 0.03 percentage points. The mean fertilization rate was 72.3%.

#### 2.3.5 Downstream biological transitions

For stages not tracked by HFEA, we synthesized evidence from published studies totaling 435,291 additional observations:

##### Blastocyst formation

Age-specific rates from Romanski et al. [23] (3,362 patients) and Sainte-Rose et al. [24] (4,952 zygotes) show stable conversion (60-67%) through age 40 with decline thereafter. We modeled this using n-weighted logistic regression with natural splines (3 degrees of freedom) to capture the age-specific relationship.

##### Euploidy

Large PGT-A datasets from Franasiak et al. [25] (15,169 biopsies) and Armstrong et al. [26] (86,208 cycles) reveal a characteristic pattern: relatively stable rates in the 20s, gradual decline through the 30s, and acceleration after age 35. We applied n-weighted logistic regression with natural splines (2 degrees of freedom) to capture this non-linear relationship.

##### Vitrification survival

Contemporary vitrification achieves 89.1% oocyte survival [29] and 95% blastocyst survival [27]. For PGT-A cycles, biopsy and testing turnaround necessitate freeze-all with deferred transfer [35].

##### Live birth after euploid transfer

While HFEA tracks live births, only 1% of UK cycles utilize PGT-A [36], necessitating external sources for euploid-specific outcomes. We analyzed SART national registry data [37] to capture clinic-level variation in euploid transfer success rates. Recognizing substantial heterogeneity across clinics, we stratified 72 qualified clinics (those with ≥100 total transfers and ≥20 transfers per age group) into performance tiers based on weighted average live birth rates. The top 10% of clinics (8 clinics, 23,324 transfers) achieved markedly higher success rates than the bottom 10% (8 clinics, 17,542 transfers), with the full dataset comprising 325,600 transfers across all qualified clinics. We trained separate n-weighted logistic regression models for each tier (top 10%, average, bottom 10%), enabling predictions that account for clinic quality. These tier-specific euploid transfer rates (e.g., 63% at age 35 for top clinics versus 42% for bottom clinics) substantially exceed unscreened HFEA rates, consistent with established benefits of euploid screening [36].

#### 2.3.6 Frozen egg integration

For frozen egg cycles, patients enter their egg count as a known value at the mature egg stage, creating a point mass distribution with downstream stages experiencing probabilistic outcomes. When no frozen egg count is specified, the cycle is modeled as a standard fresh retrieval using the age at freezing. The multi-cycle framework allows combining frozen and fresh cycles (Appendix A.1).

### 2.4 Validation Strategy

All models were validated using stratified 70/30 train-test splits by age group. Performance metrics (R^2^, RMSE, MAE) are reported on held-out test sets to ensure generalization (Appendix A.3.2). Overfitting was assessed via train-test performance gaps. Additionally, distributional predictions were evaluated using proper scoring rules (Appendix A.3.1), calibration metrics for prediction intervals (50%, 80%, 90% coverage), and probability integral transform (PIT) analysis for distributional calibration.

## 3 Results

### 3.1 Technical Validation

The model validation demonstrates excellent out-of-sample performance with minimal overfitting. Train-test gaps are under 0.007 for all models, indicating robust generalization. Beyond traditional *R*^2^ metrics, we evaluated the distributional predictions using calibration analysis. The simplified egg retrieval model achieved well-calibrated prediction intervals, with actual coverage rates closely matching nominal levels (50% coverage: 50.2%, 80% coverage: 79.2%, 95% coverage: 94.9%). These technical validation metrics confirm the model’s reliability before examining clinical applications.

### 3.2 Age-Stratified Model Predictions

To understand the model’s predictions across different age groups, we generated age-stratified outcomes for a standard clinical scenario.

These age-stratified outcomes reveal substantial within-age variability across all stages. The 95% confidence intervals span 10–45 eggs at all ages, illustrating why uncertainty quantification is essential. For example, a 35-year-old could retrieve anywhere from 3 to 42 eggs, ultimately leading to 0–14 euploid embryos. This wide range underscores the limitations of point estimates and motivates our distributional approach.

### 3.3 Clinical Application

#### 3.3.1 Single-Cycle Predictions

To demonstrate how the model applies to individual patients, we present detailed predictions for a representative clinical scenario: a 35-year-old woman with an elective baseline fertility profile undergoing stimulated ICSI in the U.S.-calibrated setting at a top 10% clinic. This example illustrates the complete distributional output at each biological stage.

The model yields an eggs-retrieved distribution with mean 17.3, median 16, and 95% interval [3, 42]. The implied probability bins are: *P* (0 eggs) = 1.0%, *P* (1–10) = 26.4%, *P* (11–20) = 40.7%, and *P* (*>* 20) = 32.0%. Propagating through the biological pipeline, the euploid-embryo distribution has mean 5.1, median 4, and 95% interval [0, 14], with *P* (0) = 4.9%, *P* (1–3) = 33.2%, and *P* (≥4) = 61.9%. The final live-birth count distribution has mean 3.0, median 3, and 95% interval [0, 9], with *P* (0) = 11.6%, *P* (1) = 18.0%, *P* (2) = 19.0%, and *P* (*≥* 3) = 51.4%. The probability of at least one live birth is 88.4%.

#### 3.3.2 Dynamic Updating with Known Values

A key innovation of our framework is the ability to update predictions as treatment progresses. To demonstrate this capability, we examined how predictions change for our 35-year-old patient when different egg retrieval outcomes are observed.

Before treatment begins, the patient expects 5.1 euploid embryos (95% CI: 0–14) with *P* (≥1 live birth) = 88.4% and *P* (≥5 euploid embryos) = 49.0%. When actual retrieval outcomes become known, all downstream predictions update accordingly (Figure 2):

**Figure 2.**
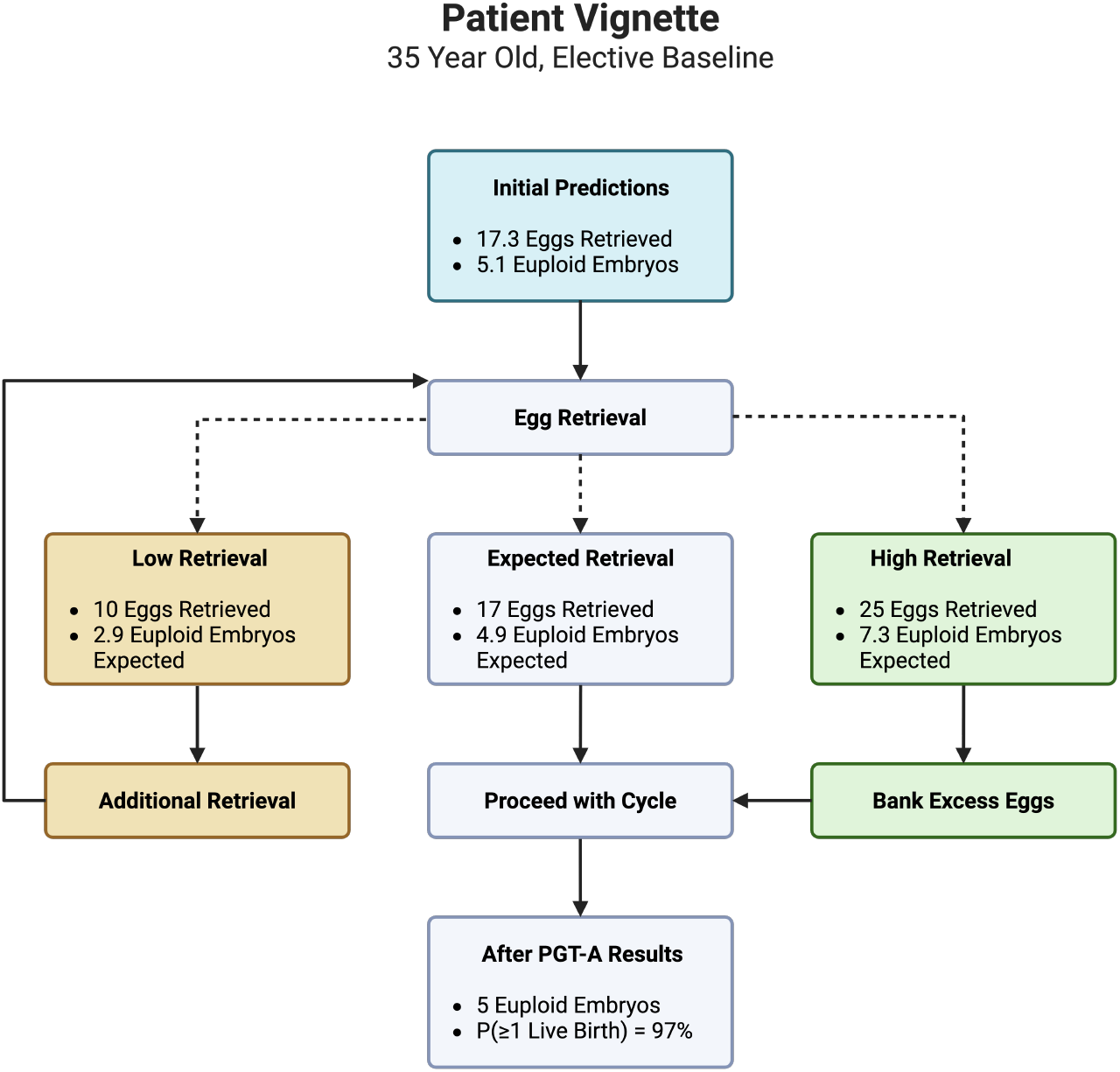
Clinical vignette demonstrating dynamic prediction updating for a 35-year-old woman with elective baseline fertility profile. Initial predictions (17.3 eggs, 5.1 euploid embryos) update based on actual retrieval outcomes, with downstream predictions and clinical recommendations adjusting accordingly. The framework enables evidence-based counseling at each decision point.

##### Low retrieval scenario (10 eggs)

Expected euploid embryos drop to 2.9 (95% CI: 0–6), *P* (≥1 live birth) = 85.0%, and *P* (≥5 euploid embryos) = 13.8%. The confidence interval width reduces by 57%, reflecting decreased uncertainty once the retrieval outcome is known.

##### Expected retrieval scenario (17 eggs confirmed)

Expected euploid embryos remain at 5.0 (95% CI: 2–9), *P* (≥1 live birth) = 96.0%, and *P* (≥5 euploid embryos) = 59.0%. The confidence interval width reduces by 50% compared to baseline.

##### High retrieval scenario (25 eggs)

Expected euploid embryos increase to 7.3 (95% CI: 3–12), *P* (≥1 live birth) = 99.0%, and *P* (≥5 euploid embryos) = 90.0%. Despite the higher count, confidence interval width still reduces by 36%.

These results demonstrate how observed outcomes narrow uncertainty while updating expectations proportionally to the deviation from baseline predictions.

#### 3.3.3 Multi-Cycle Planning

Many patients undergo multiple IVF cycles or combine fresh cycles with previously frozen gametes. Our framework handles these complex scenarios through distribution convolution while maintaining cycle-specific biology.

Consider our example patient who froze 20 eggs at age 32 and now, at age 35, contemplates a fresh cycle. The model predicts her frozen eggs will yield 6.6 euploid embryos (95% CI: 3–12), benefiting from the younger age’s higher euploidy rates. A fresh cycle at her current age would add 5.1 euploid embryos (95% CI: 0–14). Combined, she expects 11.7 euploid embryos total, substantially increasing her probability of achieving larger family size goals. Critically, the model maintains age-specific success rates rather than averaging: eggs from age 32 retain their 76% euploidy rate while those from age 35 experience 71% euploidy, demonstrating why cycle-specific modeling matters for accurate multi-cycle predictions.

## 4 Discussion

Existing IVF calculators provide either cumulative success rates (CDC, OPIS) or stage-specific point estimates (Orchid), but none quantify uncertainty or adapt to observed outcomes during treatment. We developed a distributional framework that propagates full probability distributions through each biological stage and updates predictions when actual outcomes become known. Unlike static calculators, our predictions update with observed outcomes.

The distributional approach reveals critical insights for clinical counseling. A 35-year-old woman pursuing elective embryo freezing expects 5.1 euploid embryos, but the 95% CI of 0– 14 communicates the substantial uncertainty that point estimates mask. As demonstrated in our Results, when this patient retrieves only 10 eggs, her probability of achieving five or more euploid embryos, a common threshold for meaningful genetic screening, drops from 49.0% to 13.8%. This dramatic shift provides quantitative justification for discussing additional cycles. Conversely, retrieving 25 eggs increases this probability to 90.0%, offering reassurance that costly additional procedures may be unnecessary. These evidence-based updates align counseling with each patient’s actual biological response.

Our choice of interpretable models over black-box approaches follows recent evidence from reproductive medicine AI. Complex neural networks offer marginal improvements over logistic regression for IVF prediction while sacrificing clinical interpretability [38]. The need for transparency in reproductive AI has been widely recognized, as embryo screening algorithms must be “glass-box” rather than opaque to maintain clinical trust [39]. Our framework achieves both goals: train-test gaps under 0.007 demonstrate robust generalization while preserving complete transparency in how predictions change with each variable. This interpretability proves essential when clinicians must explain why a patient’s chances improved or declined based on observed outcomes.

Beyond single-cycle predictions, multi-cycle planning represents another advance enabled by distribution convolution. When combining 20 frozen eggs from age 32 with a fresh cycle at age 35, the model maintains age-specific euploidy rates (76% vs 71%) rather than averaging. This biological accuracy matters: incorrectly pooling ages would overestimate older patients’ chances while underestimating younger patients’ outcomes. The framework thus supports complex family planning scenarios increasingly common with elective preservation, where patients bank eggs across multiple ages before attempting pregnancy. This cycle-specific approach preserves the uncertainty inherent in combining independent attempts, with variance appropriately increasing for multi-cycle predictions.

The integration of 539,215 observations provides statistical power unavailable to single-center models. While recent work shows center-specific machine learning models can outperform national registry-based predictions [19], such approaches require extensive local data unavailable to most clinics. Our framework balances generalizability with specificity through the fertility profile system, capturing clinically meaningful heterogeneity without requiring clinic-level retraining. The regional calibration factors acknowledge systematic protocol differences between U.S. and UK practices while maintaining the underlying biological relationships.

Several design choices warrant discussion. The HFEA data itself imposed several constraints: grouped reporting necessitated midpoint imputation, potentially biasing extreme values, while the absence of key predictors (BMI, AMH, AFC) limited our variable selection. Additionally, our model assumes a single infertility diagnosis per patient, as the HFEA data contains insufficient samples for each combination of multiple diagnoses to reliably model their interactions, despite some patients presenting with multiple contributing factors.

The framework’s emphasis on uncertainty quantification addresses a fundamental challenge in fertility counseling. Studies consistently show patients struggle to interpret probabilistic information and systematically overestimate their immediate success chances [11, 12]. By providing confidence intervals and tail probabilities alongside expected values, clinicians can better convey both realistic expectations and the inherent variability of biological processes. The dynamic updating feature facilitates shared decision-making by demonstrating how observed outcomes shift probabilities.

## 5 Conclusions

This distributional framework with conditional updating advances IVF prediction beyond static point estimates to dynamic, uncertainty-aware guidance. By maintaining full probability distributions and updating predictions with observed outcomes, clinicians can provide personalized counseling that evolves throughout treatment. The model’s interpretability, validated on 539,215 observations with minimal overfitting, ensures both clinical trust and robust performance. As precision medicine transforms reproductive care, tools that quantify uncertainty while remaining clinically interpretable will prove essential for optimal patient outcomes. The model is integrated as a web application at https://www.herasight.com/ivf-calculator.

## Data Availability

HFEA data are available online at https://www.hfea.gov.uk/about-us/data-research/. The IVF calculator developed in this study is publicly available at https://www.herasight.com/ivf-calculator. The underlying data is not publicly available.

https://www.herasight.com/ivf-calculator

## A Supplementary Methods

### A.1 Technical Details of Multi-Cycle Modeling

#### A.1.1 Zero-Inflated Negative Binomial Model for Egg Retrieval

The egg retrieval model uses a zero-inflated negative binomial (ZINB) distribution to accommodate both excess zeros (cancelled cycles, poor responders) and overdispersion in count data. The probability mass function is:

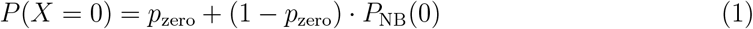

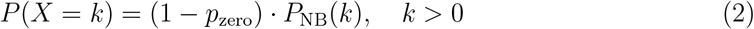

where *P*_NB_(*k*) denotes the negative binomial probability mass function with parameters *µ* (mean) and *θ* (dispersion), parameterized as:

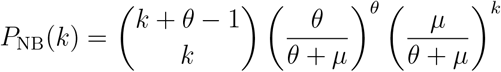

The zero-inflation probability *p*_zero_ and mean *µ* are modeled as functions of patient covariates through logit and log links respectively.

#### A.1.2 Discrete Convolution for Cycle Combination

When combining independent IVF cycles, the total count distribution follows the convolution of individual distributions:

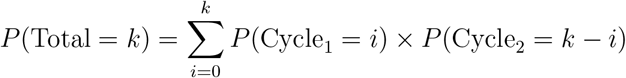

For computational efficiency, the implementation uses Fast Fourier Transform (FFT) based convolution when distribution sizes exceed 100 elements, reducing complexity from O(*n*^2^) to O(*n* log *n*). When per-stage or per-cycle success probabilities differ, the resulting total follows a Poisson-binomial distribution [30]. This operation preserves the mathematical properties of independent random variables:

- *E*[Total] = *E*[Cycle_1_] + *E*[Cycle_2_]
- Var(Total) = Var(Cycle_1_) + Var(Cycle_2_)

To maintain computational tractability, distributions exceeding 500 elements are truncated while preserving 99.9999% of the probability mass. This truncation retains the values with the highest probability density, typically centered around the mode, ensuring minimal information loss. However, distributions derived from point masses (known values) bypass truncation to maintain precision.

#### A.1.3 Point Distributions from Known Values

When a known value is specified at any stage in a cycle, that stage’s distribution becomes:

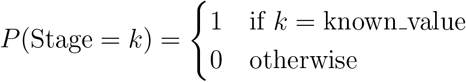

This point distribution propagates through downstream stages via binomial filters, creating appropriate variance for biological processes while maintaining the deterministic starting point. For frozen egg cycles, the known value is entered at the mature egg stage.

### A.2 Model Coefficients

The following table presents coefficient estimates from all regression models used in our framework. All models were trained on the 70% training subset of HFEA data and validated on the 30% held-out test set. Coefficients represent log-odds changes for binomial models (maturity, fertilization) and log-count changes for the zero-inflated negative binomial model (egg retrieval). Statistical significance is assessed at conventional levels (*p <* 0.05).

The table compares simplified (parsimonious) and full (non-parsimonious) model specifications. The simplified models retain only predictors with meaningful impact on held-out test performance, achieving test *R*^2^ values within 0.2 percentage points of the full models while requiring fewer user inputs. The full models include all available predictors for transparency, though the marginal improvements in test *R*^2^ (1.6–2.0 percentage points for egg retrieval, negligible for others) do not justify the additional complexity.

**Table A1:**
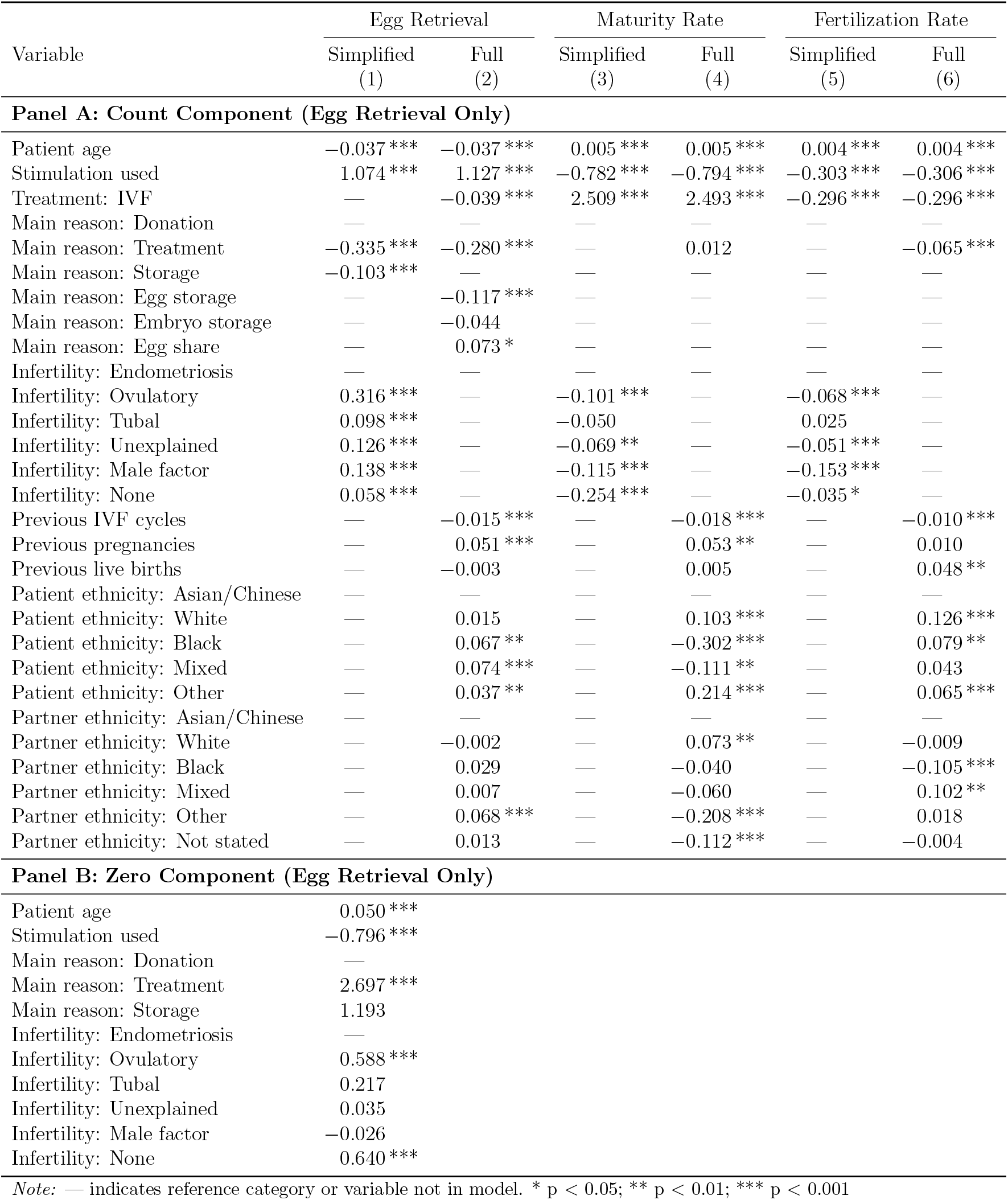
Model Coefficients: Simplified and Full Specifications.

| Variable | Egg Retrieval |  | Maturity Rate |  | Fertilization Rate |  |
| --- | --- | --- | --- | --- | --- | --- |
|  | Simplified<br>(1) | Full<br>(2) | Simplified<br>(3) | Full<br>(4) | Simplified<br>(5) | Full<br>(6) |
| **Panel A: Count Component (Egg Retrieval Only)** |  |  |  |  |  |  |
| Patient age | −0.037 *** | −0.037 *** | 0.005 *** | 0.005 *** | 0.004 *** | 0.004 *** |
| Stimulation used | 1.074 *** | 1.127 *** | −0.782 *** | −0.794 *** | −0.303 *** | −0.306 *** |
| Treatment: IVF | — | −0.039 *** | 2.509 *** | 2.493 *** | −0.296 *** | −0.296 *** |
| Main reason: Donation | — | — | — | — | — | — |
| Main reason: Treatment | −0.335 *** | −0.280 *** | — | 0.012 | — | −0.065 *** |
| Main reason: Storage | −0.103 *** | — | — | — | — | — |
| Main reason: Egg storage | — | −0.117 *** | — | — | — | — |
| Main reason: Embryo storage | — | −0.044 | — | — | — | — |
| Main reason: Egg share | — | 0.073 * | — | — | — | — |
| Infertility: Endometriosis | — | — | — | — | — | — |
| Infertility: Ovulatory | 0.316 *** | — | −0.101 *** | — | −0.068 *** | — |
| Infertility: Tubal | 0.098 *** | — | −0.050 | — | 0.025 | — |
| Infertility: Unexplained | 0.126 *** | — | −0.069 ** | — | −0.051 *** | — |
| Infertility: Male factor | 0.138 *** | — | −0.115 *** | — | −0.153 *** | — |
| Infertility: None | 0.058 *** | — | −0.254 *** | — | −0.035 * | — |
| Previous IVF cycles | — | −0.015 *** | — | −0.018 *** | — | −0.010 *** |
| Previous pregnancies | — | 0.051 *** | — | 0.053 ** | — | 0.010 |
| Previous live births | — | −0.003 | — | 0.005 | — | 0.048 ** |
| Patient ethnicity: Asian/Chinese | — | — | — | — | — | — |
| Patient ethnicity: White | — | 0.015 | — | 0.103 *** | — | 0.126 *** |
| Patient ethnicity: Black | — | 0.067 ** | — | −0.302 *** | — | 0.079 ** |
| Patient ethnicity: Mixed | — | 0.074 *** | — | −0.111 ** | — | 0.043 |
| Patient ethnicity: Other | — | 0.037 ** | — | 0.214 *** | — | 0.065 *** |
| Partner ethnicity: Asian/Chinese | — | — | — | — | — | — |
| Partner ethnicity: White | — | −0.002 | — | 0.073 ** | — | −0.009 |
| Partner ethnicity: Black | — | 0.029 | — | −0.040 | — | −0.105 *** |
| Partner ethnicity: Mixed | — | 0.007 | — | −0.060 | — | 0.102 ** |
| Partner ethnicity: Other | — | 0.068 *** | — | −0.208 *** | — | 0.018 |
| Partner ethnicity: Not stated | — | 0.013 | — | −0.112 *** | — | −0.004 |
| **Panel B: Zero Component (Egg Retrieval Only)** |  |  |  |  |  |  |
| Patient age | 0.050 *** |  |  |  |  |  |
| Stimulation used | −0.796 *** |  |  |  |  |  |
| Main reason: Donation | — |  |  |  |  |  |
| Main reason: Treatment | 2.697 *** |  |  |  |  |  |
| Main reason: Storage | 1.193 |  |  |  |  |  |
| Infertility: Endometriosis | — |  |  |  |  |  |
| Infertility: Ovulatory | 0.588 *** |  |  |  |  |  |
| Infertility: Tubal | 0.217 |  |  |  |  |  |
| Infertility: Unexplained | 0.035 |  |  |  |  |  |
| Infertility: Male factor | −0.026 |  |  |  |  |  |
| Infertility: None | 0.640 *** |  |  |  |  |  |

*Note:* — indicates reference category or variable not in model. \* p < 0.05; \*\* p < 0.01; \*\*\* p < 0.001

### A.3 Technical Validation Metrics

While the main paper focuses on clinically interpretable metrics, we also evaluated our distributional predictions using standard probabilistic forecasting metrics for methodological completeness.

#### A.3.1 Proper Scoring Rules

We assessed the quality of our probabilistic predictions using two proper scoring rules:

- **Log Score**: Measures the logarithmic probability assigned to the observed outcome. The mean log score across test observations was 3.7, indicating the model assigns reasonable probability mass to actual outcomes.
- **Continuous Ranked Probability Score (CRPS)**: Quantifies the integrated squared difference between predicted and observed cumulative distributions. The mean CRPS was 5.1, suggesting good overall distributional accuracy.

These metrics, while standard in probabilistic forecasting literature, lack direct clinical interpretation and thus are reported here for technical validation rather than in the main results.

#### A.3.2 Model Performance (R-squared)

To quantify predictive performance, we evaluated both simplified and full model specifications using coefficient of determination (R^2^) on held-out test sets. As shown in the Results section (Table 1), the simplified models achieve test R^2^ values of 0.137 for egg retrieval, 0.879 for maturity rate, and 0.643 for fertilization rate.

**Table 1:** Out-of-sample model performance: Simplified vs Full models with train-test validation.

| Model | Test $R^2$ | | Train $R^2$ | | Train-Test Gap (Full) |
| --- | --- | --- | --- | --- | --- |
|  | Simplified | Full | Simplified | Full |  |
| Egg Retrieval | 0.137 | 0.153 | 0.140 | 0.155 | 0.002 |
| Maturity Rate | 0.879 | 0.879 | 0.886 | 0.886 | 0.007 |
| Fertilization Rate | 0.643 | 0.643 | 0.648 | 0.649 | 0.005 |

**Table 2:** Age-stratified predictions (mean [95% interval]) for elective baseline fertility profile, ICSI, stimulation, U.S. calibration, and top 10% clinic performance.

| Age | Eggs retrieved | Mature eggs | Fertilized | Blastocysts | Euploid embryos |
| --- | --- | --- | --- | --- | --- |
| 30 | 22.1 [4–52] | 18.1 [3–43] | 13.0 [2–32] | 8.8 [1–22] | 6.7 [0–17] |
| 35 | 17.3 [3–42] | 14.3 [2–35] | 10.3 [1–25] | 7.1 [0–18] | 5.1 [0–14] |
| 40 | 13.9 [2–34] | 11.5 [1–28] | 8.3 [1–21] | 5.6 [0–15] | 2.9 [0–8] |
| 42 | 12.5 [1–31] | 10.4 [1–26] | 7.5 [0–19] | 4.4 [0–12] | 1.8 [0–6] |

These results demonstrate that:

- The egg retrieval model shows meaningful improvement (1.6 percentage points) from the full specification.
- Maturity and fertilization models show negligible improvements from additional predictors.
- All models exhibit minimal overfitting with train-test gaps under 0.007.
- The high R^2^ values for maturity (0.879) and fertilization (0.643) rates indicate strong predictive performance for these binary outcomes when converted from rate to count predictions.

The minimal train-test gaps validate our model selection approach and confirm that the reported performance metrics reflect true generalization capability rather than in-sample fitting.

